# Groundbreaking predictions about COVID-19 pandemic duration, number of infected and dead: A novel mathematical approach never used in epidemiology

**DOI:** 10.1101/2020.08.05.20168781

**Authors:** J. G. García de Alcañíz, V. López-Rodas, E. Costas

## Abstract

Hundreds of predictions about the duration of the pandemic and the number of infected and dead have been carried out using traditional epidemiological tools (i.e. SIR, SIRD models, etc.) or new procedures of big-data analysis. However, the extraordinary complexity of the disease and the lack of knowledge about the pandemic (i.e. R value, mortality rate, etc.) create uncertainty about the accuracy of these estimates. However, several elegant mathematical approaches, based on physics and probability principles, like the Delta-*t* argument, Lindy’s Law or the Doomsday principle-Carter’s catastrophe, which have been successfully applied by scientists to unravel complex phenomena characterized by their great uncertainty (i.e. Human race’s longevity; How many more humans will be born before extinction) allow predicting parameters of the Covid-19 pandemic. These models predict that the COVID-19 pandemic will hit us until at least September-October 2021, but will likely last until January-September 2022, causing a minimum of 36,000,000 infected and most likely 60,000,000, as well as 1,400,000 dead at best and most likely 2,333,000.

Competing Interest Statement

The authors have declared no competing interest.

## Introduction

The sudden arrival of a new and unknown virus^1^ has unleashed a global pandemic which all countries are still fighting but with very different results. This is not the only problem with SARS-CoV-2, the uncertainty about the evolution of the COVID-19 pandemic is colossal.

Scientists are making a tremendous effort to understand and counteract the effect of the virus. Up to date, more than 7000 preprints are available for researches, many focus on clinical aspects of the disease^2–14^, others on virus’ features, some more on epidemiological characteristics, etc. When confronted with unknown events, mathematical approaches are very useful delivering new knowledge rapidly^15–18^.

From the very beginning, numerous mathematical approaches have proven their usefulness to better understand COVID-19 outbreak.

Regression analysis have shed light into aspects of the disease that may allow governments to make better decisions, generally the first useful mathematical approach^2,19–22^. Some country variables like tourism, mobility and pollution predict well the number of infected and dead, whereas national health system, economic status, etc. predict to a much lesser grade^23^

Other mathematical approach to the COVID-19 problem is epidemiology using traditional tools or the more recent big-data analysis. Many traditional predictive models about the duration of the pandemic or about the number of infected have been carried out using epidemiology numerical tools (i.e. SIR, SIRD models^24–26^, Gompertz’s equation^27–32^, etc., but, as good as these tools may be, some factors contribute to create uncertainty about the usefulness of these traditional models with COVID-19 outbreak:

- Lack of knowledge about the disease or SARS-CoV-2 itself (i.e. key parameters to typify the evolution of the pandemic: *R* value, infectivity rate mortality rate, etc.).
- The extraordinary complexity of a disease that has spread around the globe and is challenging different countries with different climates, different wealth, different social structure, with very different strategies to control the pandemic, etc.
- Reliability of the official data (false or biased data, different methodologies to register cases or deaths among countries, etc.). Official figures of infected or dead do not match those obtained by serology tests (i.e. Carlos III Health Institute study)^33^.

These factors hamper the relevance and reliability of the traditional epidemiological models.

In addition, due to some uncertainties that arise derived from the lack of knowledge, predictions about the pandemic are unreliable. Some of these crucial unanswered questions could be: How long will the natural immunity last after overcoming the disease? Will there be interactions or synergies with influenza virus when the season comes? Will SARS-CoV-2 jump back to animals (domestic or wildlife) and will these species act as natural reservoirs? Will people comply with health authorities’ policies? Will there be effective vaccines or drugs? When will all this happen?

However, there are some elegant mathematical approaches, based on basic science, physics and probability principles, like the Copernican principle and the Delta-*t* argument, Lindy’s Law, the Doomsday principle-Carter’s catastrophe, all of which allow predicting complex phenomena characterized by their great uncertainty, as the Covid-19 pandemic is. These mathematical procedures have been successfully applied by scientists to unravel intricate problems (i.e. Human race’s longevity; How many more humans will be born before extinction?) or to more mundane problems (i.e. predict in the 60’s when the berlin wall will fall, how long a Broadway musical will be on show or how long will it take for a company to shut down).

The surprisingly effective predictive power of these approaches made us apply them to estimate how long COVID-19 will last, how many people in total will be infected and how many will dye, as well as a final distribution of infected and dead in the different countries. There is no other paper in the literature exercising these other successfully effective mathematical tools.

We will do two different approaches:

1. Probabilistic calculations based on the Copernican principle that will allow us to define:
  1.1. Probable duration of the pandemic using Delta-*t* argument ^34,35^.
  1.2. Probable duration of the COVID-19 pandemic based on Lindy’s law^36^.
2. Probabilistic calculations based on Doomsday argument that will enable us to define:
  2.1. Total number of infected and dead with the Doomsday argument^37,38^

SARS-CoV-2 has challenged mankind. Under the best scenario our model predicts, the COVID-19 pandemic will hit us until, at least, September-October 2021 (it will likely last until January-September 2022), causing a minimum of 36,000,000 infected (most likely 60,000,000), and 1,400,000 dead (most likely 2,333,000).

Theoretical background

### 1. Probable duration of the Covid-19 pandemic

#### 1.1. Estimating total duration of the COVID-19 pandemic with Delta-*t* argument

Applying the Copernican principle (earth does not occupy, nor in space nor in time, a privileged position in the universe) to the study of diverse scientific phenomenon has allowed notable progress in science^34,35^. The fact that for any given event at any moment in time (i.e. up to date COVID-19 pandemic) there are no privileged observers, no special moments, allows robust duration predictions.

Assuming that any event we observe can only be measured between initial time (*t_begin_*) and final time (*t_end_*) and that we are non-privileged observers of such event, then current time (*t_now_*) will be randomly placed in any possible moment throughout the duration of the event. In such way, ratio *r* = *(t_now_ − t_begin_)/(t_end_ − t_now_)* is a random number between 0 and 1. This enables the statistical calculation of the probability of any future event (Fig. 1).

**Figure 1.**
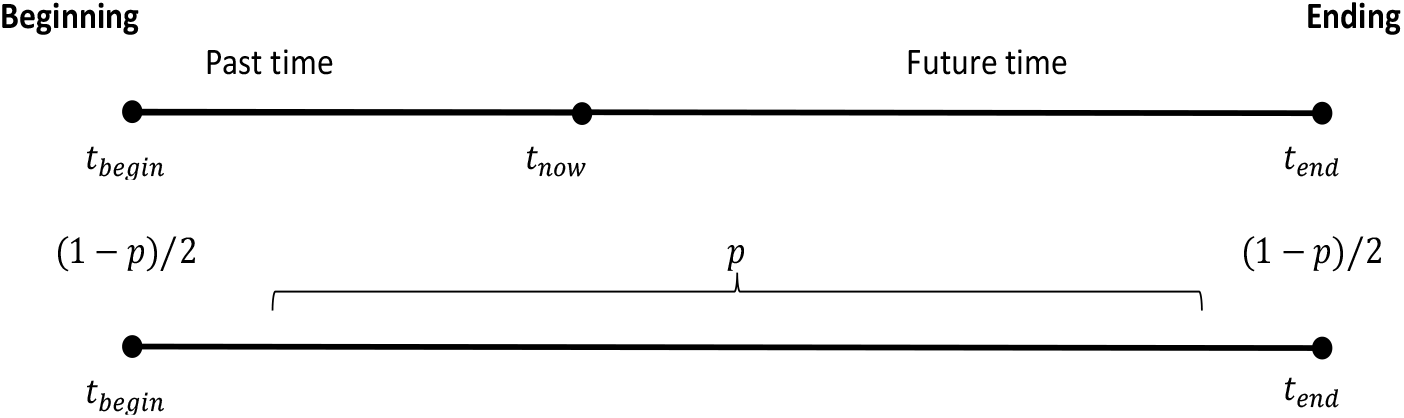
Time distribution probability of a non-privileged event

In this way

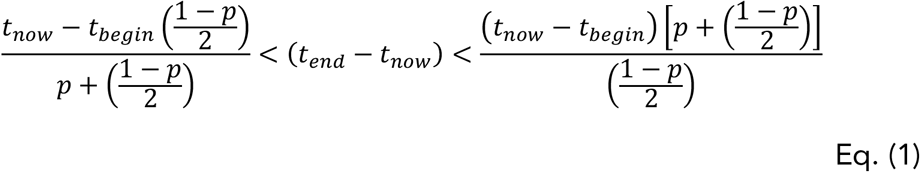

future duration of the COVID-19 pandemic can be calculated, with its related probability.

#### 1.2. Duration of the COVID-19 pandemic based on Lindy’s law

Lindy’s effect assumes that, future life expectancy of a phenomenon is proportional to its current age. Consequently, whenever Lindy’s effect applies, every additional survival period implies longer life expectancy remaining. Mathematically is described as follows:

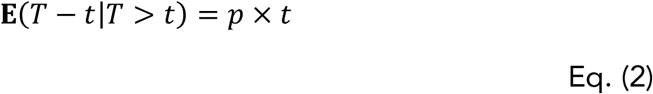

Where *T* is the random time under consideration (i.e. lifetime of the COVID-19 pandemic), which takes values in the range *(c* ≤ *t <* ∞), and *p* is the Lindy proportion, a positive parameter^36^.

In our model we estimate COVID-19 pandemic lifetime assuming three different *p* values which fit with previous pandemic outbreaks of SARS-CoV-1, H1N1 and MERS)

#### 1.3. Estimating total number of infected and dead by COVID-19 pandemic worldwide using the Doomsday argument

In 1998, J. Leslie, using the Copernican principle and based on previous works by Carter (1983) and Gott (1994), calculated the total number of people to be born before total extinction of the human race.

In the same way, we propose to use this same principle to calculate the total number of infected and dead by COVID-19.

Be *N* the total number of people to be infected by COVID-19, we will call it Future stage; and *n* the number of people infected up to date by COVID-19, named Present stage.

Assuming the Copernican principle (there is no privilege or special moment regarding COVID-19) and that we are in a nonspecial place then it abides to:

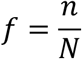

uniformly distributed in the interval (0,1]. So, at any given time for a nonspecial observer there is a probability *p* = 0.95 that *f* is in the interval (0.05,1] and then

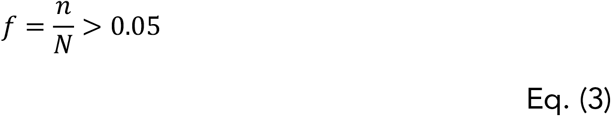

Which allows to estimate the total number of infected and dead by SARS-CoV-2.

## Results

### 1. Most probable duration of the COVID-19 pandemic

#### 1.1. Estimating COVID-19 pandemic total lifetime with Delta-*t* argument

Chinese authorities identified human cases with onset of Covid-19 symptoms in early December 2019. So, we assume for our estimations that Covid-19 pandemic has a *t_begin_ =* December 10^th^, 2019 and a *t_now_ =* August 10^th^, 2020. A summary of the observed predictions using in Eq (1) these values and three different significant levels (*p =* 0.5; *p* = 0.7; *p* = 0.95) can be found in Table 1.

**Table 1.** Estimated duration of COVID-19 pandemic based on Copernican principle (Delta-*t* argument). Type I error: The prediction indicates that the pandemic has finished while it still remains. Type II error: The prediction indicates that the pandemic remains active while it has finished.

| Median pandemic duration | Maximum pandemic duration | Minimum pandemic duration | $p$ |
| --- | --- | --- | --- |
| until October 2021 | until October 2022 | until November 2020 | $p = 0.5$<br>Increases the likelihood of Type I error |
| until September 2022 | until late November 2024 | until late September 2020 | $p = 0.7$ |
| until 2035 | until late 2049 | | $p = 0.95$<br>Increases the likelihood of Type II error |

Delta-*t* argument foresees a likelihood setting in which COVID-19 pandemic could end between second half of 2021 and the end of 2022. Looking into more optimistic settings could lead to Type I errors (predict a future date with no pandemic when in fact still will be). Predictions that COVID-19 pandemic will last beyond 2024 could fall into Type II errors (predict a future date with pandemic when it will have already ended).

#### 1.2. Estimating COVID-19 lifetime with Lindy’s law

Table 2 shows an estimate of the COVID-19 pandemic duration assuming a t value of nine months (*t_begin_* =December 10^th^, 2019 and a *t_now_* =August 10^th^, 2020) and three different values for *p*, Lindy’s proportion.

According to Lindy’s effect, COVID-19 pandemic should end between September 2021 and October 2022.

**Table 2.** Predictions about duration of COVID-19 pandemic based on Lindy’s effect. Type I error: The prediction indicates that the pandemic has finished while it still remains. Type II error: The prediction indicates that the pandemic remains active while it has finished.

| Pandemic duration ($T$) | Lindy's proportion ($p$) |
| --- | --- |
| Until September 2021 | 0.5<br>Increases the likelihood of<br>Type I error |
| Until January 2022 | 1 |
| Until October 2022 | 2<br>Increases the likelihood of<br>Type II error |

### 2. Estimating worldwide total number of infected and dead by COVID-19 pandemic using The Doomsday Argument

Knowing that at the present position (July 31^st^, 2020) there are worldwide 17,064,064 confirmed cases (https://covid19.who.int) and using Eq. (3), the maximum number of infected by SARS-CoV-2 should always be under 360,000,000 people (*p* = 0.95).

Similarly, knowing that at the same position in time there are 668,073 deaths (https://covid19.who.int) the estimate of deaths should always be under 14 million people (*p* = 0.95).

Results are presented on Table 3.

**Table 3.** Predictions about number of infected and dead by COVID-19 based on the Doomsday argument. (Calculations based on FECHA data: 18,000,000 infected and 700,000 dead) Type I error: the prediction indicates a fewer number of infected and dead than will occur. Type II error: the prediction indicates a higher number of infected and dead than will occur.

| Total number of infected by the end of the COVID-19 pandemic | Total number of dead by the end of the COVID-19 pandemic | *p* |
| --- | --- | --- |
| 36,000,000 | 1,400,000 | 0.5<br>Increases the likelihood of Type I error |
| 60,000,000 | 2,333,100 | 0.70 |
| 360,000,000 | 14,000,000 | 0.95<br>Increases the likelihood of Type II error |

## Discussion

Newton’s hope, reflected in his *Principia Mathematica*, was that knowing present conditions and the instantaneous rate of change, future could be accurately predicted^39^. However, since Gödel’s incompleteness theorems^40^ demonstrating the inherent limitations of every formal axiomatic system capable of modelling basic arithmetic, it was evident that science would have certain limitations to achieve accuracy in Newtonian prediction^41^.

The geological instant in which we are living is characterized by increasing global extinction rates, homogenization of biotas, proliferation of opportunistic species and pest-and-weed ecology, all of which favor the occurrence of unpredictable emergent novelties (reviewed by Myers and Knoll 2001 and Woodruff 2001)^42,43^. COVID-19 pandemic can be a very good example of an unpredictable emergent novelty, its arrival was particularly difficult to predict. For example, for the turn of the century, Oxford University Press gathered in a book what 30 of the brightest minds of the time thought the future would bring. None predicted a pandemic would devastate the world^44^.

SARS-CoV-2 took us by surprise and we underestimated it, but once its real and true magnitude was clear hundreds of scientists started modeling to predict both duration in time and number of infected and dead. Epidemiology has an important theoretical body and, up to date, different predictive models have been used^45–55^.

Some problems arise with these COVID-19 epidemiology models because they make complex assumptions due to lack of information about key aspects like number of cases, transmission rates, contact parameters, immunity; how they display uncertainty in the model; which data is being used; is it general or focuses on a particular setting; etc.^56–59^, these assumptions hinder the accuracy of the predictions. Predictive models at a global scale, even at country level are more inaccurate than a local scale^59^.

According to Holmdhal and Buckee three model parameters in particular limit our ability to predict the future of the Covid-19: the extent of protective immunity, the extent of transmission and immunity among asymptomatic people or with minimal symptoms and the measurements of contact rates between susceptible and infectious people^56^.

Unlike traditional models, the assumptions we propose are much simpler. Models based on the Delta-*t* argument and Lindy’s law only need duration of the pandemic until present date and Doomsday argument models need the number of infected and dead, which makes predictions at a global scale easier. But, are the predictions these models produce correct?

Both, model based on Delta-*t* argument and model based on Lindy’s law predict that COVID-19 pandemic will last, at the very least, until September-October 2021 and probably will remain until same period of 2022. Model based on the Doomsday argument predicts a minimum of 36 million people infected and 1.4 million dead, but the most probable figures count up to 60 million infected and 2.333 million dead. It is worth noting, that these three different models produce predictions consistent with each other.

In this sense, expected pandemic duration using Delta-*t* argument or Lindy’s effect fits in what is reasonable according to what epidemiologists predict using traditional models. Similarly, infected and dead estimate using the Doomsday model look also rational. Up to date different models have predicted a wide range duration of the COVID-19 pandemic.

However, the onset of contingent events impossible to predict and very unlikely, the Black Swan effect^60^ have great influence over the accuracy of these models based on the Copernican principle (i.e. Delta-*t* argument or Doomsday argument). Contingent events, like the emergence of mutations that significantly vary the infectivity or the case fatality rate of the SARS-CoV-2, developing a vaccine that can be massively administered, finding of an effective drug used worldwide could significantly alter our predictions. After all, any prediction based on the Copernican principle will be true only if, and only if, “nor the studied phenomenon nor the observer occupy no special position in space or time”. It is also possible that there are so many factors that intervene in the evolution of the COVID-19 pandemic that any prediction is scientifically indefinable.

There have been refutations to the Doomsday and Delta-*t* arguments at a theoretical level, mainly coming from philosophy and psychology areas^61–63^.

But the success predicting very different events, from the fall of the Berlin wall predicted on the 60’, the demise of the U.S.S.R. in the 70’ to the longevity of Broadway musicals give present work more relevance to understand what may happen if we do not find soon a reliable pharmaceutical solution to SARS-CoV-2.

## Data Availability

All data was obtained from public web pages and links are available in the manuscript

